# A Clinical Predictor of Lung Molecular Endotype Identifies Heterogeneity in Corticosteroid Response in Severe COVID-19: an Emulated Target Trial

**DOI:** 10.64898/2026.06.08.26355201

**Authors:** Benjamin J Sines, Robert S Hagan, Xi Jiang, Ella Pavlechko, Scott McClain, Xin Hunt, Julia Florou-Moreno, Jake Acquadro, Gabriel Risa, Varunraj Valsaraj, Jonathan C Schisler, Matthew C Wolfgang

## Abstract

**Background:** Corticosteroids reduce mortality in severe COVID-19 requiring oxygen or invasive mechanical ventilation, yet emerging data suggest that SARS-CoV-2-associated acute lung injury is biologically heterogeneous and that treatment response may vary across molecularly defined disease states. Lung-derived molecular endotypes of severe COVID-19-associated acute lung injury have been described, but direct molecular profiling is not routinely available at the bedside. We evaluated whether a clinical predictor of previously defined lung molecular endotype identifies heterogeneity in corticosteroid treatment effect among mechanically ventilated patients with COVID-19.

**Methods:** We utilized a single-center cohort of 5,000 patients with COVID-19 treated at the University of North Carolina Hospital between January 1, 2020, and December 31, 2022, to emulate a target trial assessing the effect of corticosteroid receipt on mortality, length of stay, and incident organ support. Confounding was addressed through inverse probability of treatment weighting (IPTW). Outcomes for severely ill patients requiring mechanical ventilation were compared to the RECOVERY trial results, with subsequent moderation analysis and stratified analysis by clinically predicted lung molecular endotype and vaccination status. The primary outcome was 28-day mortality. Secondary Outcomes were time to discharge alive and progression to additional organ support.

**Results:** This emulated target trial showed a directionally favorable but non-statistically significant association between corticosteroid treatment and reduced 28-day mortality in patients requiring mechanical ventilation for SARS-CoV-2 infection. A clinical predictor of lung molecular endotype moderated the effect of corticosteroids on 28-day mortality (p-value for interaction 0.038) and identified distinct predicted endotype-specific treatment effect. Corticosteroid treatment was associated with lower 28-day mortality in the predicted Hyper-Inflammatory endotype (OR 0.62, 95% CI 0.39, 0.99) but not in the predicted Metabolic Dysregulation endotype (OR 1.15, 95% CI 0.82, 1.61). We did not detect significant effect modification by vaccination status (p-value for interaction 0.65), although inference was limited by the small, vaccinated subgroup (28-mortality OR 0.78, 95% CI 0.37, 1.65 in vaccinated vs 0.94, 95% CI 0.70, 1.26 in unvaccinated).

**Conclusions:** In this target trial emulation of mechanically ventilated patients with severe COVID-19, corticosteroid treatment showed a directionally favorable but non-statistically significant association with reduced 28-day mortality in the overall cohort. However, a clinical predictor of lung molecular endotype identified significant heterogeneity in treatment effect, with benefit concentrated in the predicted Hyper-Inflammatory endotype and no apparent benefit in the predicted Metabolic Dysregulation endotype. These findings support prospective validation of clinically deployable endotype-guided corticosteroid treatment strategies in acute lung injury and ARDS.

## BACKGROUND

The RECOVERY and REMAP-CAP trials provide evidence supporting corticosteroid administration for severe COVID-19 requiring hospitalization and supplemental oxygen.^1,2^ These trials were conducted largely before widespread vaccination and before later SARS-CoV-2 variants, leaving important questions about the consistency of corticosteroid benefit across evolving populations of critically ill patients. In RECOVERY, the positive effect of corticosteroids was observed only in the subset of patients requiring supplemental oxygen. Within this subset, those requiring mechanical ventilation demonstrated greater benefit than those requiring supplemental oxygen alone. Despite the significant mortality benefit observed in the RECOVERY trial, the statistical endpoint for superiority was not met in the REMAP-CAP trial, where a Bayesian analysis suggested a 93% probability of superiority of steroids compared with standard care.^2^ Subsequent analyses demonstrate heterogeneous treatment effect in the subpopulation of patients with acute respiratory distress syndrome (ARDS) using post-hoc molecularly defined clusters of hyper- and hypo-inflammatory ARDS.^3,4^ However, molecularly defined latent classes are not currently available at the bedside to guide real-time treatment decisions.

Emerging evidence suggests that the molecular pathologic drivers of acute lung injury from SARS-CoV-2 infection are heterogeneous.^4–7^ Most of these analyses have focused on risk stratification^8^, severity of disease in hospitalized patients^9,10^, or mortality outcomes.^1^ Additionally, the clinical manifestation of COVID-19 is heterogeneous.^11,12^ Exploratory analysis suggests that a previously defined hyperinflammatory phenotype of ARDS is less common in COVID-19-associated ARDS as compared to ARDS from other etiologies.^7^ In non-randomized retrospective analysis, a hyperinflammatory phenotype demonstrated improved mortality with receipt of corticosteroids and convalescent plasma, and a hypoinflammatory phenotype demonstrated worsened mortality with corticosteroid receipt.^4,13,14^ Together, these observations support the concept that average treatment effects may obscure biologically meaningful subgroups with divergent responses to immunomodulatory therapy.

We previously identified two lung-derived molecular endotypes of severe COVID-19-associated acute lung injury using tracheal aspirate immune mediator and transcriptomic profiling. These endotypes are referred to here as the Hyper-Inflammatory endotype and the Metabolic Dysregulation endotype. The Hyper-inflammatory endotype demonstrated molecular features consistent with corticosteroid responsiveness, whereas the Metabolic Dysregulation endotype showed features suggesting reduced corticosteroid responsiveness. Because direct airway molecular profiling is not routinely available at the time of treatment assignment, a clinically deployable predictor of lung endotype could provide a practical approach to endotype-informed corticosteroid treatment.

Clinical decision making informed by underlying molecular disturbances, rather than poorly calibrated clinical severity metrics such as oxygen requirement alone, may improve allocation of available interventions, concentrating benefit and avoiding harms.^15–17^ Molecularly informed clinical decision aids in ARDS are still exploratory. Additionally, the effect of vaccination status on ARDS pathobiology and response to corticosteroids in the subpopulation of patients with COVID-19-associated ARDS has not been fully evaluated.

Although corticosteroids are guideline-concordant for severe COVID-19 and are now conditionally recommended for ARDS more broadly, they are not benign.^18,19^ In critically ill patients, corticosteroids may contribute to hyperglycemia, impaired wound healing, and secondary bacterial or fungal infections.^20,21^ These risks may be amplified in patients requiring mechanical ventilation, invasive devices, and prolonged immobility.^20^ Thus, identifying patients most likely to benefit from corticosteroids could increase therapeutic value while reducing avoidable exposure among likely non-responders.

This study addresses ongoing questions regarding the universal benefit of corticosteroid receipt in a critically ill and mechanically ventilated population of patients with COVID-19-associated ARDS. We utilize a cohort that spans SARS-CoV-2 epochs (alpha, delta, omicron) and vaccination periods to first benchmark corticosteroid effect on 28-day mortality in mechanically ventilated patients with COVID-19 against a mechanically ventilated subgroup of the RECOVERY trial. We subsequently evaluate heterogeneity of treatment effect across a clinical predictor of lung molecular endotype and vaccination status.

## METHODS

### Data Sources

The retrospective cohort was derived from inpatient admissions at the University of North Carolina at Chapel Hill Hospital between January 1, 2020, and December 31, 2022. This study was performed in accordance with the principles of the Declaration of Helsinki, and institutional review board approval was obtained from the University of North Carolina Office of Human Research Ethics before the start of the study (IRB study number 22-3196, date of approval: 1/11/2023).

The University of North Carolina Health Care System catalogues clinical data from the electronic medical record, research data, and administrative data in the Carolina Data Warehouse for Health for archival and research purposes. Electronic medical record data is stored for all encounters and across data domains in the PCORnet common data model, capturing clinical data from the current electronic medical record back to April 2014. All affiliated hospitals utilizing this medical record are included in the Carolina Data Warehouse database. All analyses follow the Strengthening the Reporting of Observational Studies in Epidemiology (STROBE) guidelines and the instrument to assess the credibility of effect modification analyses (ICEMAN) framework for effect modification analysis.^22,23^

### Cohort Construction

The target trial emulation criteria are outlined in **Supplementary Table 1**. We identified all adults (age >=18) admitted to the University of North Carolina Hospitals with a new oxygen requirement and SARS-CoV-2 positive testing on admission by PCR between January 1, 2020, and December 31, 2022. We limited the study population to individuals requiring mechanical ventilation within 7 days of admission. Time zero was defined as initiation of invasive mechanical ventilation. Active treatment included those receiving a glucocorticoid equivalent to 6 mg of dexamethasone (**Supplementary Table 1**) within 48 hours of initiation of mechanical ventilation and was compared to those who did not receive corticosteroids in this time interval. This 48-hour grace period was selected to approximate clinical treatment assignment at the time of mechanical ventilation while allowing for real-world variation in medication administration timing. The sample timeline of a patient assigned to the treatment arm is shown in **Supplementary Figure 1**.

Study subjects with contraindications to corticosteroids or severe immune deficiencies (active cytotoxic therapy, solid organ or bone marrow transplant recipients, acute leukemia, chronic corticosteroid use) were excluded from analysis, following the RECOVERY Trial protocol. Patients with another clear indication for corticosteroids, including COPD exacerbation, adrenal insufficiency, or severe shock, were also excluded when identifiable from the electronic medical record.

### Analytic Framework

We employed a target trial emulation framework for causal inference to assess the effect of corticosteroids versus supportive care alone on 28-day mortality in patients hospitalized and requiring mechanical ventilation for ARDS associated with COVID-19 (**Supplementary Table 1**).

Secondary outcomes include 28-day incident requirements for additional organ support (new requirement of vasopressors or renal replacement therapy after treatment assignment) and time to discharge alive. Consistent with the intention to treat analytic framework, untreated study participants who crossed over to receive treatment after the eligibility time frame were not censored and instead analyzed as untreated. All subjects were followed through day 28 and censored if they were transferred prior to that time to a different inpatient acute care hospital. Patients discharged to hospice were considered equivalent to discharged as deceased. All other discharges prior to day 28 were assumed alive through day 28 and cross-referenced with separate mortality tables.

We balanced baseline covariates by utilizing the inverse probability of a study subject receiving their assigned treatment. Weights were derived from logistic regression models adjusting for age, sex, race, history of chronic obstructive lung disease, history of cardiovascular disease, CKD stages 3-5, diabetes, and renal replacement therapy prior to enrollment.

### IPTW diagnostics

The mean inverse probability of treatment weight (IPTW) across the cohort was equal to 2.0, and the standard deviation of the inverse probability of treatment weights was equal to 0.99. The minimum and maximum inverse probability of treatment weights were 1.0 and 6.9 respectively. There was no evidence of non-positivity or misspecification of the propensity score models based on examination of the distributions of weights. Baseline covariates in this matched cohort were compared to assess for balance with a threshold of absolute standardized mean difference (SMD) <0.10 (or 10%) indicating balance between the groups. The Love plot in **Supplementary Figure 3** shows the improvement in balance in the weighted population by absolute standardized mean difference.

### A priori stratification

We have previously defined two discrete molecular endotypes of severe COVID-19 based on lung rather than blood sampling.^24^ These molecular classifications were derived from immune effector and transcriptomic analysis from tracheal aspirate samples in mechanically ventilated patients with severe COVID-19 as well as SARS-CoV-2 negative controls. For this study, the previously defined endotypes are referred to as the Hyper-Inflammatory endotype and the Metabolic Dysregulation endotype. The Hyper-inflammatory endotype was hypothesized a priori to be the corticosteroid-responsive endotype based on prior lung molecular profiling.

We applied a machine learning model following the predictive modeling protocol published and described previously.^25^ The model predicted endotype class assignment for subjects lacking molecular data, enabling assessment of heterogeneous treatment effect of corticosteroids across predicted molecular sub-populations using 24-hour trends in respiratory rate, albumin, mean corpuscular hemoglobin, and anion gap. The predictions were based on clinical features extracted from clinical data collected within 48 hours before the patient’s first requirement for mechanical ventilation, as well as historical data from the medical record, including premorbid diagnoses, condition, and medication administration history prior to ventilation, all processed according to the previously described feature engineering pipeline.^26^ The model output was a probability of assignment to the Hyper-inflammatory endotype. Predicted endotype was dichotomized using a prespecified probability threshold of 0.5, with probabilities greater than 0.5 assigned to the Hyper-inflammatory endotype and probabilities less than or equal to 0.5 assigned to the Metabolic Dysregulation endotype.

Vaccination status was defined as receipt of at least one SARS-CoV-2 directed vaccine 14 days or greater prior to diagnosis with COVID-19. All comorbidities were defined by ICD10 codes available from the electronic medical record. Ages were defined at time of enrollment. Race and ethnicity were defined by self-identification or identification by next of kin when the study subject was unable to interact at the time of hospital intake.

### Heterogeneous Treatment Effect Analysis

Weighted logistic regression was performed after assignment of IPT weights to evaluate effect of corticosteroid receipt on all pre-specified outcomes. Models incorporating treatment and the pre-specified stratification variables as interaction terms were generated to evaluate the moderating effect of these variables on the effect of treatment on 28-day mortality. Stratified models were generated across predicted lung molecular endotype and vaccination status to evaluate directionality and magnitude of treatment effect. Finally, time-to-event analyses were generated on the weighted population and in lung endotype and vaccine stratified populations.

## RESULTS

Amongst 5,000 admissions with respiratory viral testing positive for SARS-CoV-2 at our center in the specified timeframe, 593 went on to require mechanical ventilation within 7 days of hospitalization. Of these, 164 were excluded based on trial exclusion criteria, including patients who were status post bone marrow or solid organ transplant, had an inherited immune deficiency, or were receiving cytotoxic or immunosuppressive treatments. Because some exclusion criteria overlapped, exclusion categories are shown in **Figure 1**. Of the original cohort, 429 were eligible and included in the emulated target trial (**Figure 1**).

**Figure 1:**
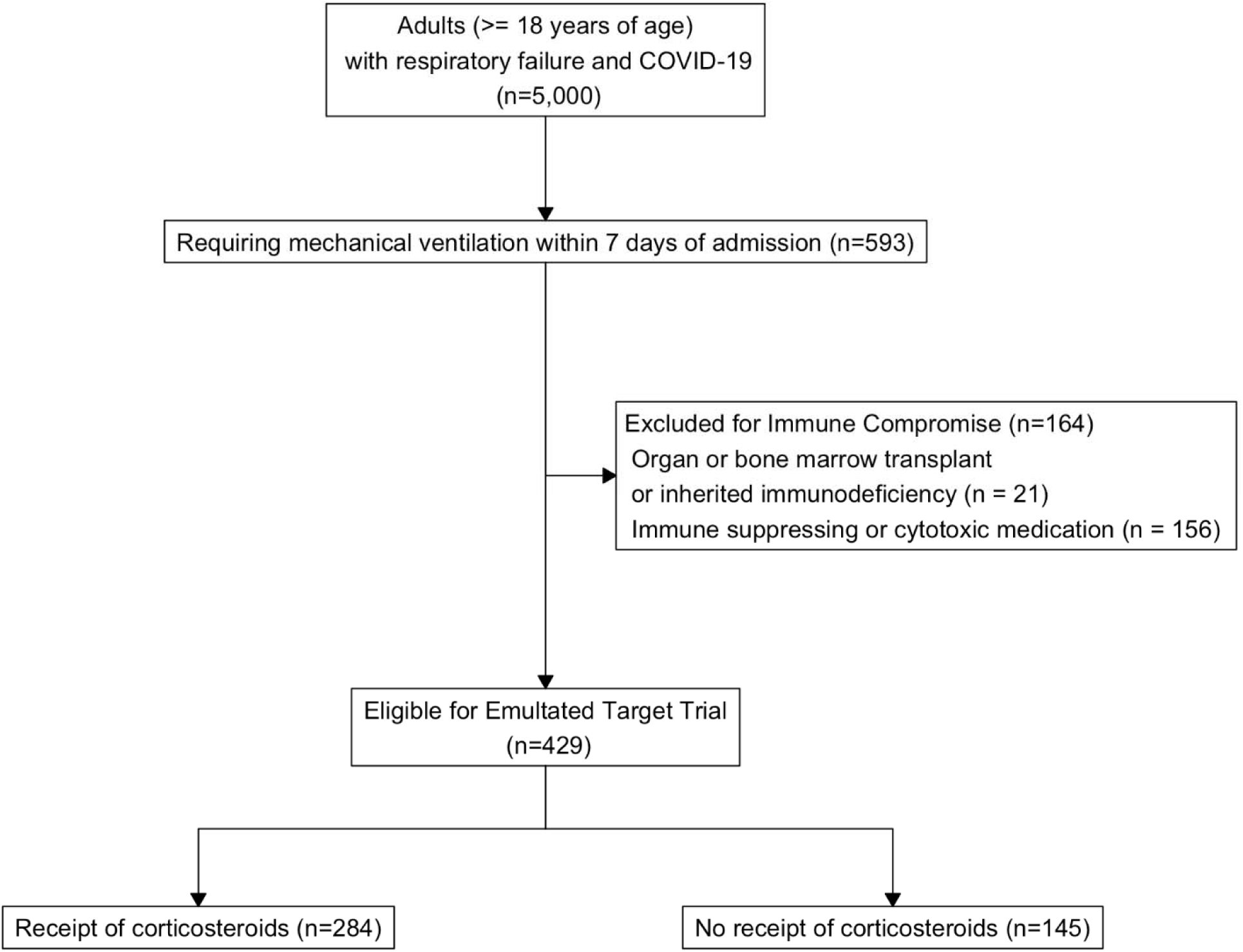
Identification of eligible emulated target trial participants

The balance of demographic and clinical characteristics of patients who received corticosteroids versus usual care alone is shown in **Table 1** and are largely balanced except for older age in recipients of corticosteroids (62 years of age versus 58.5 years of age), higher prevalence of diabetes (39.4% versus 28.3%), and higher prevalence of heart disease (52.8% versus 38.6%). When stratified by a machine learning model predictor of lung molecular endotype, this cohort was well balanced across demographic and clinical characteristics apart from a small increase in prevalence of liver disease in the predicted Metabolic Dysregulation endotype (13.4% versus 6.8%) (**Table 2**).

**Table 1:**
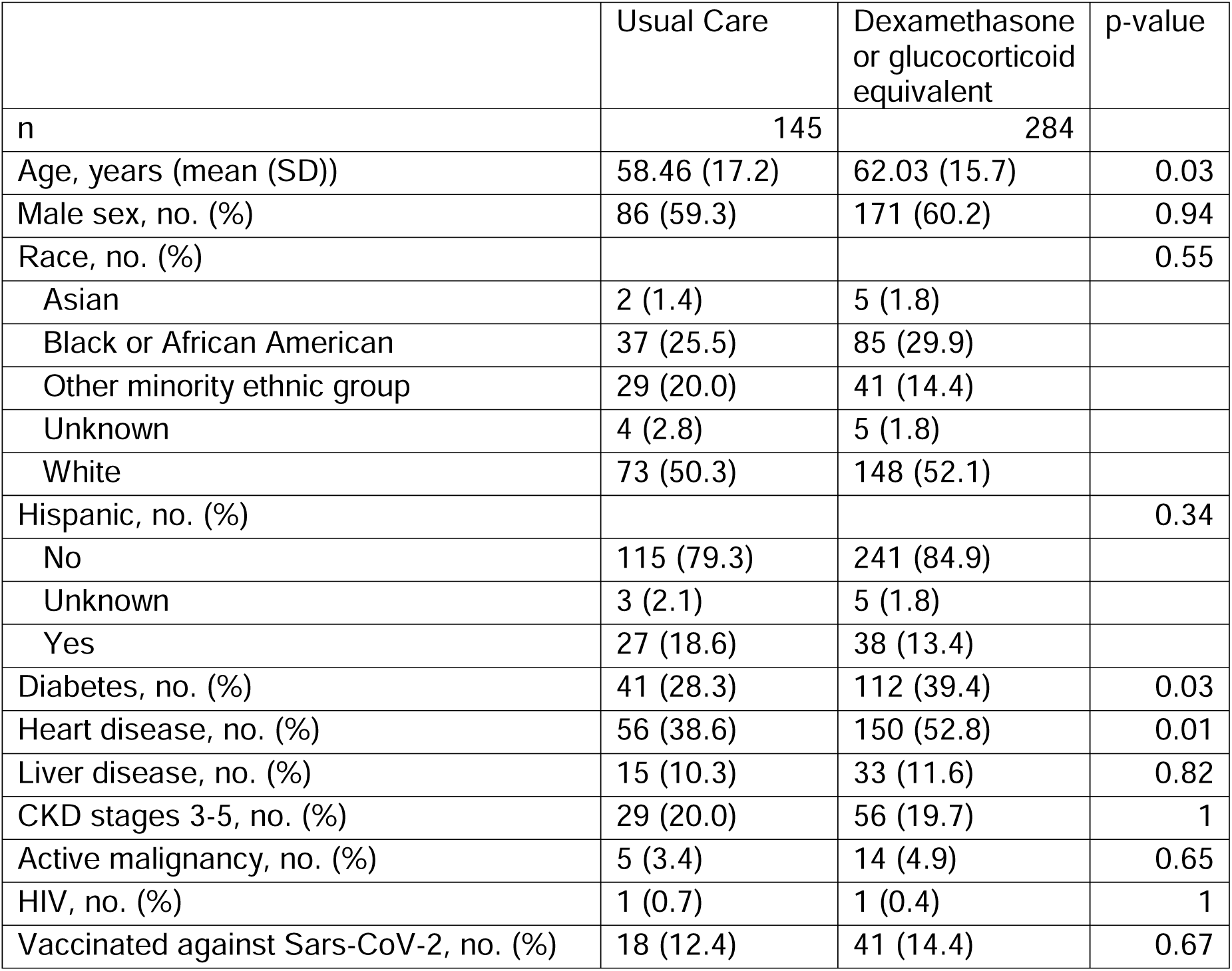
Characteristics of Patients at Baseline, by treatment receipt.

|  | Usual Care | Dexamethasone<br>or glucocorticoid<br>equivalent | p-value |
| --- | --- | --- | --- |
| n | 145 | 284 |  |
| Age, years (mean (SD)) | 58.46 (17.2) | 62.03 (15.7) | 0.03 |
| Male sex, no. (%) | 86 (59.3) | 171 (60.2) | 0.94 |
| Race, no. (%) |  |  | 0.55 |
| Asian | 2 (1.4) | 5 (1.8) |  |
| Black or African American | 37 (25.5) | 85 (29.9) |  |
| Other minority ethnic group | 29 (20.0) | 41 (14.4) |  |
| Unknown | 4 (2.8) | 5 (1.8) |  |
| White | 73 (50.3) | 148 (52.1) |  |
| Hispanic, no. (%) |  |  | 0.34 |
| No | 115 (79.3) | 241 (84.9) |  |
| Unknown | 3 (2.1) | 5 (1.8) |  |
| Yes | 27 (18.6) | 38 (13.4) |  |
| Diabetes, no. (%) | 41 (28.3) | 112 (39.4) | 0.03 |
| Heart disease, no. (%) | 56 (38.6) | 150 (52.8) | 0.01 |
| Liver disease, no. (%) | 15 (10.3) | 33 (11.6) | 0.82 |
| CKD stages 3-5, no. (%) | 29 (20.0) | 56 (19.7) | 1 |
| Active malignancy, no. (%) | 5 (3.4) | 14 (4.9) | 0.65 |
| HIV, no. (%) | 1 (0.7) | 1 (0.4) | 1 |
| Vaccinated against Sars-CoV-2, no. (%) | 18 (12.4) | 41 (14.4) | 0.67 |

**Table 2:**
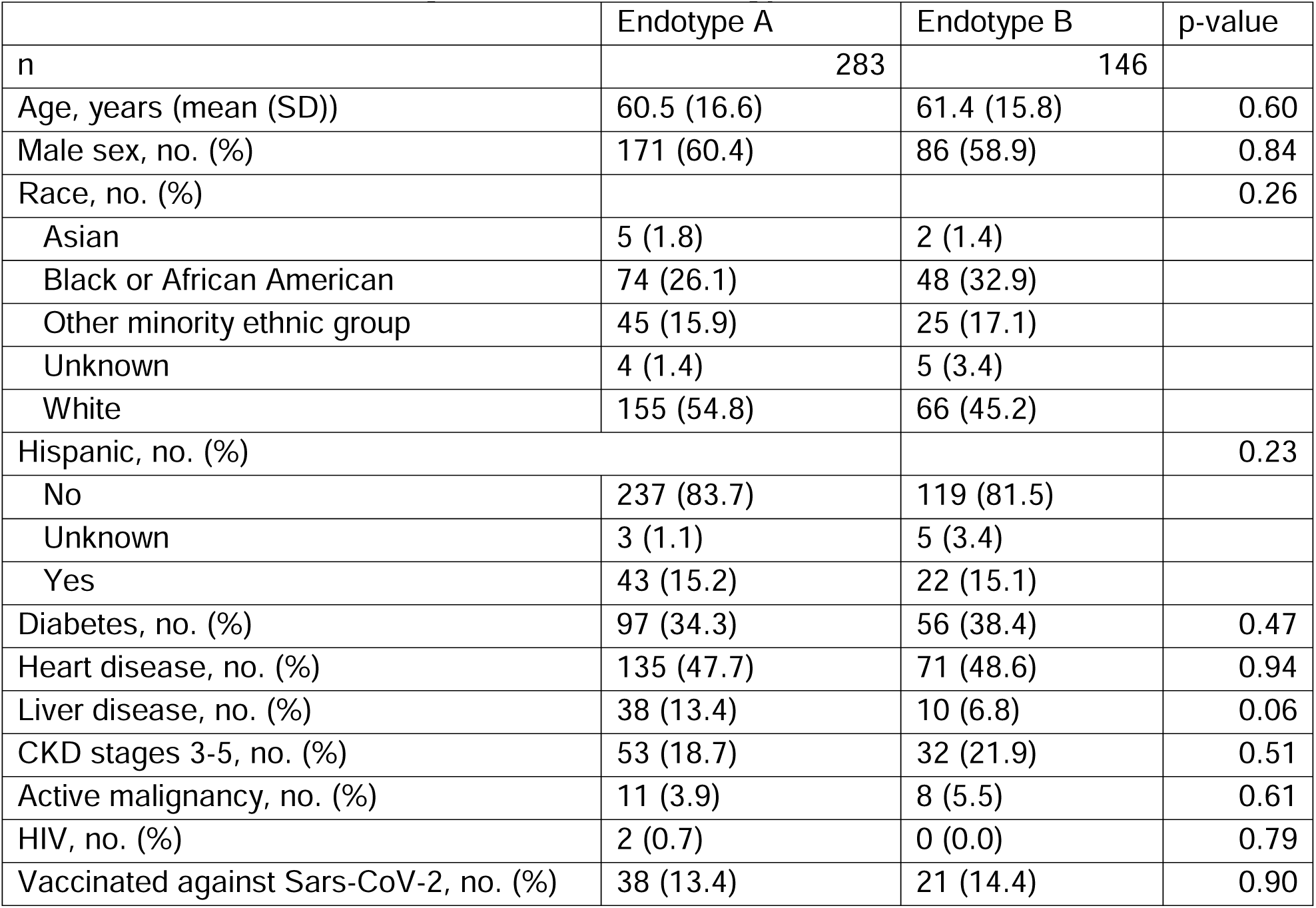
Characteristics of Cytokine Derived Endotypes.

In the unweighted and unadjusted analysis, treatment with corticosteroids was associated with 1.09 times the odds of mortality at 28 days (95% CI 0.73, 1.65). In the weighted analysis, treatment with corticosteroids at the time of initiation of mechanical ventilation was associated with lower but non-statistically significant odds of 28-day mortality (OR 0.92, 95% CI 0.70, 1.21) and a non-statistically significant lower hazard ratio (HR 0.89, 95% CI 0.64, 1.23) (**Figure 2A**).

**Figure 2:**
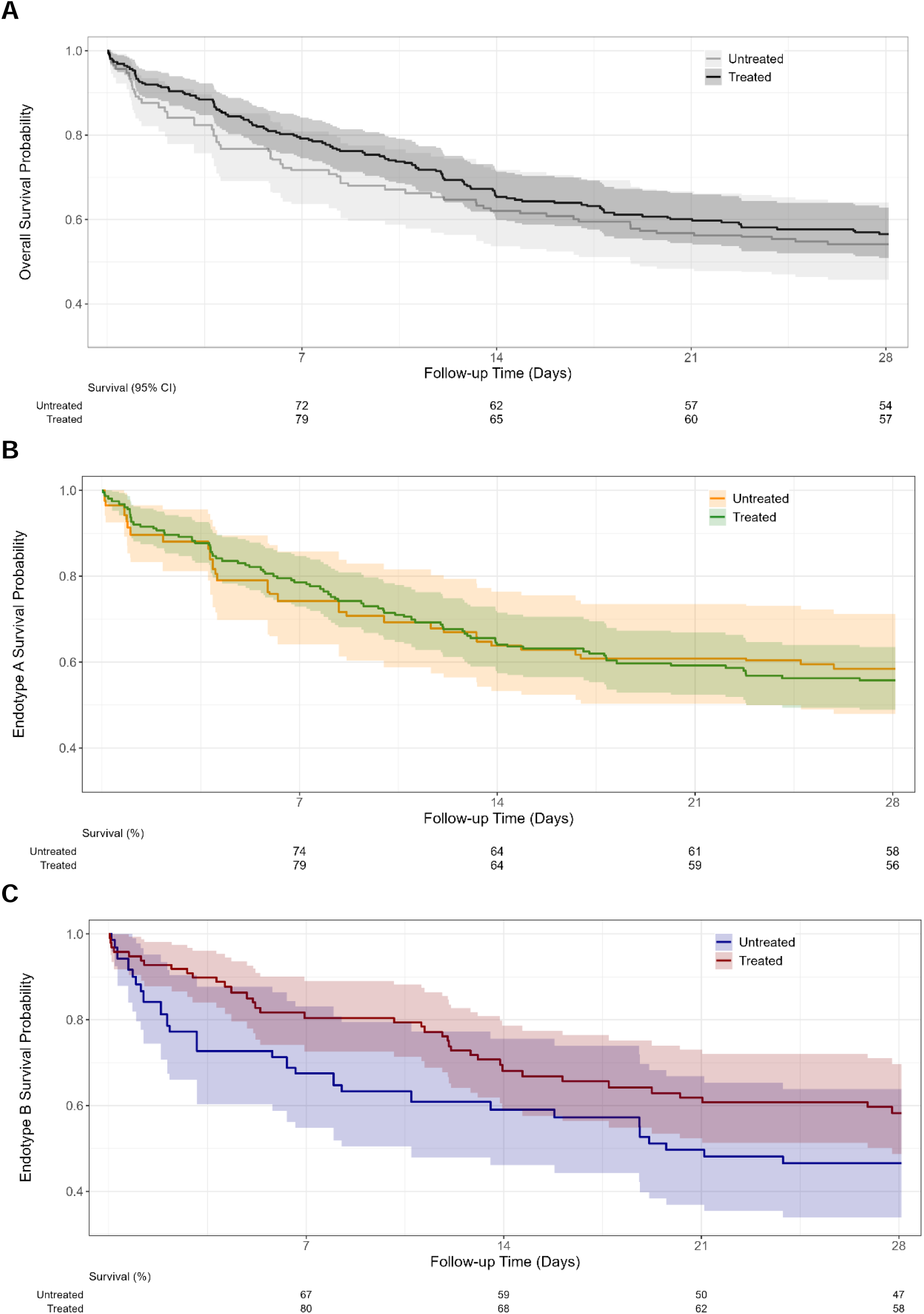
Kaplan-Meier curve for survival-mortality through day 28 in weighted population in A) full cohort (HR 0.89 [0.64, 1.23] p = 0.5), B) sub-cohort of predicted Endotype A study subjects (HR 1.04 [0.68, 1.60] p = 0.9), and C) sub-cohort of predicted Endotype B study subjects (HR 0.67 [0.4, 1.13] p = 0.14).

When stratified by clinically predicted lung endotype, 283 patients were assigned to the Metabolic Dysregulation endotype, and 146 patients were assigned to the Hyper-Inflammatory endotype. Predicted lung endotype modified the association between corticosteroid treatment and 28-day mortality (interaction p = 0.038). In weighted analyses, corticosteroid treatment was not associated with reduced 28-day mortality in the predicted Metabolic Dysregulation endotype (OR 1.15, 95% CI 0.82, 1.61), whereas corticosteroid treatment was associated with lower 28-day mortality in the predicted Hyper-Inflammatory endotype (OR 0.62, 95% CI 0.39, 0.99). Time-to-event analyses showed a similar pattern, with HR 1.04 (95% CI 0.68, 1.60) in the predicted Metabolic Dysregulation endotype and HR 0.67 (95% CI 0.40, 1.13) in the predicted Hyper-Inflammatory endotype (**Figure 2B and 2C**).

Time to discharge alive through day 28 was not significantly changed under treatment in the overall cohort nor when stratified by predicted lung molecular endotype (**Figure 3A-C**). Incident requirement for new organ support was significantly decreased for the predicted Hyper-Inflammatory endotype, with an odds ratio (OR) of 0.32 (95% CI 0.12, 0.76) for renal replacement therapy and an OR of 0.42 (95% CI 0.18, 0.94) for vasopressors. Incident requirement for renal replacement therapy alone was decreased in all groups and stratifications under treatment with corticosteroids (overall OR 0.30, p<0.0001). Incident requirement for vasopressors alone was unchanged under treatment in the overall population (OR 0.80, p=0.32) and in the predicted Metabolic Dysregulation subgroup (OR 1.05, p=0.85). Together, these secondary outcome analyses supported a broader clinical benefit signal in the predicted Hyper-Inflammatory endotype.

**Figure 3:**
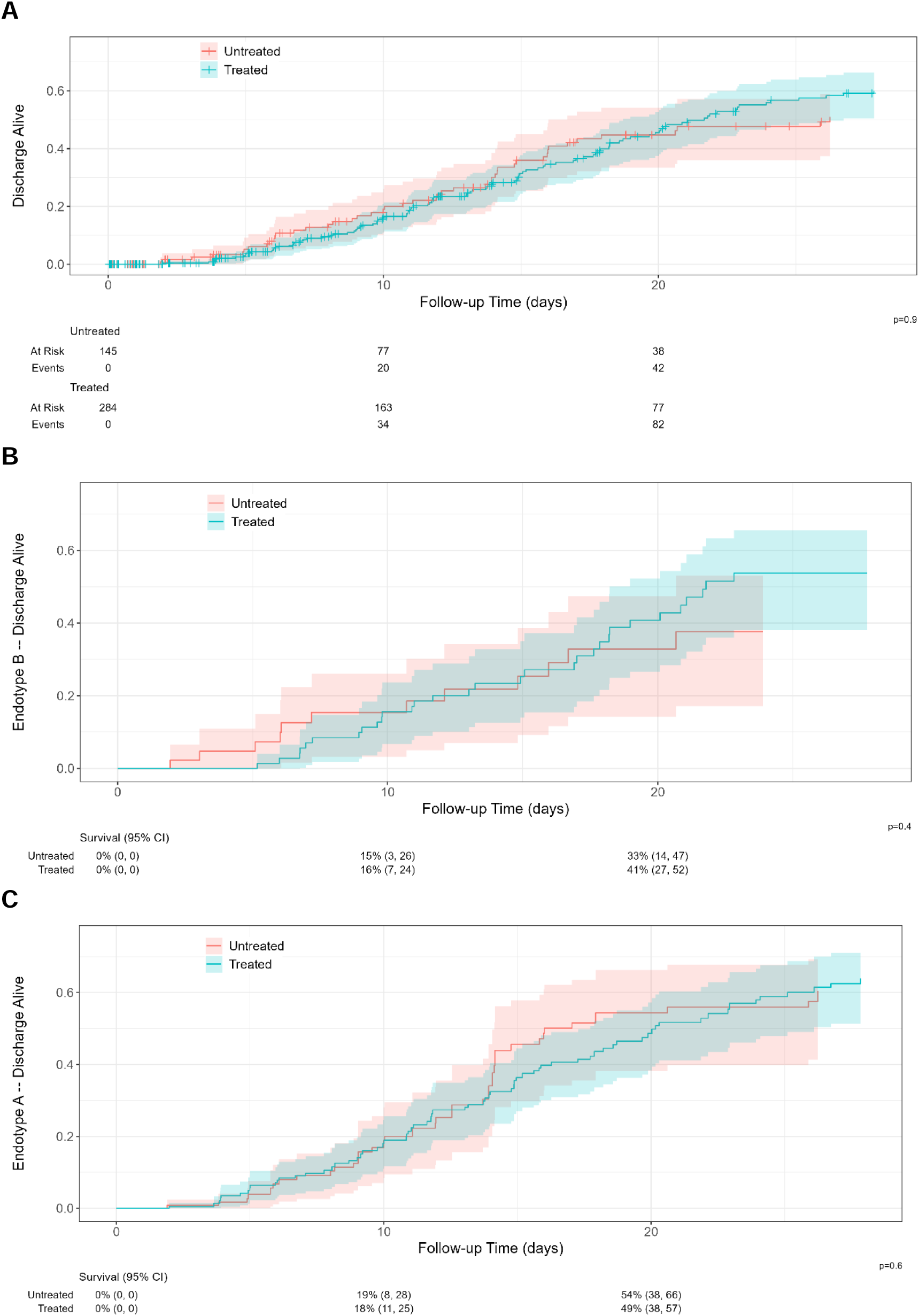
Time to discharge alive through day 28 in weighted population in A) full cohort, B) sub-cohort of predicted Endotype A study subjects, and C) sub-cohort of predicted Endotype B study subjects.

Vaccination status did not have a significant effect modification on the impact of corticosteroids on 28-day mortality, with OR 0.78 (95% CI 0.37, 1.65) and HR 0.71 (95% CI 0.31, 1.61) in the vaccinated group vs OR 0.94 (95% CI 0.70, 1.26) and HR 0.91 (95% CI 0.64, 1.30) in the unvaccinated group. Additionally, there was no difference in incident requirement for renal replacement therapy or incident requirement for vasopressors in weighted analyses across these strata. However, the vaccinated subgroup was small, limiting inference regarding vaccination as an effect modifier.

## DISCUSSION

In this target trial emulation of mechanically ventilated patients with severe COVID-19, corticosteroid treatment was associated with a directionally favorable but non-statistically significant reduction in 28-day mortality in the overall cohort. The primary finding is that this average effect obscured heterogeneity by clinically predicted lung molecular endotype. Corticosteroid benefit was concentrated in patients assigned to the predicted Hyper-Inflammatory endotype, whereas patients assigned to the predicted Metabolic Dysregulation endotype showed no evidence of benefit. These findings suggest that clinically available data may approximate lung molecular endotypes and identify subgroups of patients with differential response to immunomodulatory therapy.

The overall treatment effect observed in our cohort was more modest than that reported in the mechanically ventilated subgroup of RECOVERY. Several factors may explain this attenuation. First, this real-world cohort included patients treated across later pandemic periods, including eras with changing viral variants, vaccination availability, evolving standards of care, and shifting baseline risk. Second, patients in our cohort had high rates of prognostic comorbidities that may have diluted the observable treatment effect, even after IPTW balancing of measured baseline characteristics. Third, corticosteroid exposure was defined pragmatically as dexamethasone or glucocorticoid-equivalent treatment, which increases generalizability but may introduce heterogeneity in drug formulation, timing, and duration. Finally, the attenuated average effect may reflect true biological heterogeneity, as suggested by the endotype-stratified treatment effects observed in this analysis.

We did not detect significant effect modification by vaccination status. However, this analysis should be interpreted cautiously because the vaccinated subgroup was small and because missing vaccination data were classified as unvaccinated in the primary analysis. Thus, these findings should not be interpreted as evidence of equivalent corticosteroid benefit across vaccination strata. Rather, within this mechanically ventilated cohort, vaccination status did not identify a clear subgroup with differential corticosteroid treatment effect. Because only critically ill patients were enrolled, the independent effect of vaccination on mortality or incident organ failure could not be evaluated.

The endotype-stratified findings are consistent with the concept that acute lung injury and ARDS are biologically heterogeneous syndromes in which average treatment effects may obscure clinically relevant subgroups. The previously defined lung molecular endotypes revealed distinct immune and transcriptomic programs in severe COVID-19-associated acute lung injury. In the current study, a clinical predictor of those lung endotypes, applied using data available near the time of treatment assignment, identified patients with divergent corticosteroid treatment effects. Patients assigned to the predicted Hyper-Inflammatory endotype showed lower 28-day mortality and reduced incident organ support with corticosteroid treatment, whereas those assigned to the predicted Metabolic Dysregulation endotype did not. These data support further development of clinically deployable endotype predictors as tools for precision treatment assignment in critical illness.

This study has multiple strengths. The analysis was weighted with a well-specified IPTW procedure to balance covariates across exposed and unexposed populations. The inclusion criteria for this emulated trial mirror those of the most severely ill arm of the RECOVERY trial and are consistent with current guideline recommendations for dexamethasone while preserving a pragmatic design that increases generalizability to real-world care. The endotype analysis was based on a previously defined lung molecular classification and a previously described clinical prediction model, supporting biological plausibility and reducing the risk that the observed effect modification reflects purely post-hoc subgroup discovery.

There are some limitations of note in this analysis. Firstly, despite centralized efforts to harmonize data and support data quality, issues of data inaccuracy inherent to electronic medical records, such as degree of completeness and accuracy, cannot be fully avoided or adjusted. For example, incomplete vaccination records prevent us from perfectly determining the proportion of vaccinated patients, as missing data are assumed to indicate unvaccinated status. Additionally, incomplete medical history remains a possible concern, and a similar assumption was made in those cases. Second, we cannot exclude the possibility of residual unmeasured confounding despite the use of a well-specified IPTW procedure. Third, comparison of an active immune-suppressing intervention with supportive care invites immortal time bias, which we attempted to mitigate by tightly specifying eligibility criteria. Finally, the clinical classifier predicts, rather than directly measures, lung molecular endotype. Prospective validation with contemporaneous molecular sampling and randomized treatment assignment will be required before this approach can be implemented in clinical care.

## CONCLUSION

In conclusion, this target trial emulation found a directionally favorable but non-statistically significant association between corticosteroid treatment and reduced 28-day mortality in mechanically ventilated patients with severe COVID-19. A clinically predicted lung molecular endotype significantly modified this treatment effect, with apparent benefit concentrated in the predicted Hyper-Inflammatory endotype and no evidence of benefit in the predicted Metabolic Dysregulation endotype. These findings suggest that latent biological heterogeneity may influence the effectiveness of broadly applied, guideline-concordant immunomodulatory therapies. Prospective studies and randomized trials should evaluate whether lung endotype-guided treatment assignment can improve outcomes in acute lung injury and ARDS while limiting unnecessary corticosteroid exposure among likely non-responders.

## Data Availability

All data produced in the present study are available upon reasonable request to the authors

## Supplementary Content

STROBE Statement—Checklist of items that should be included in reports of *cohort studies*

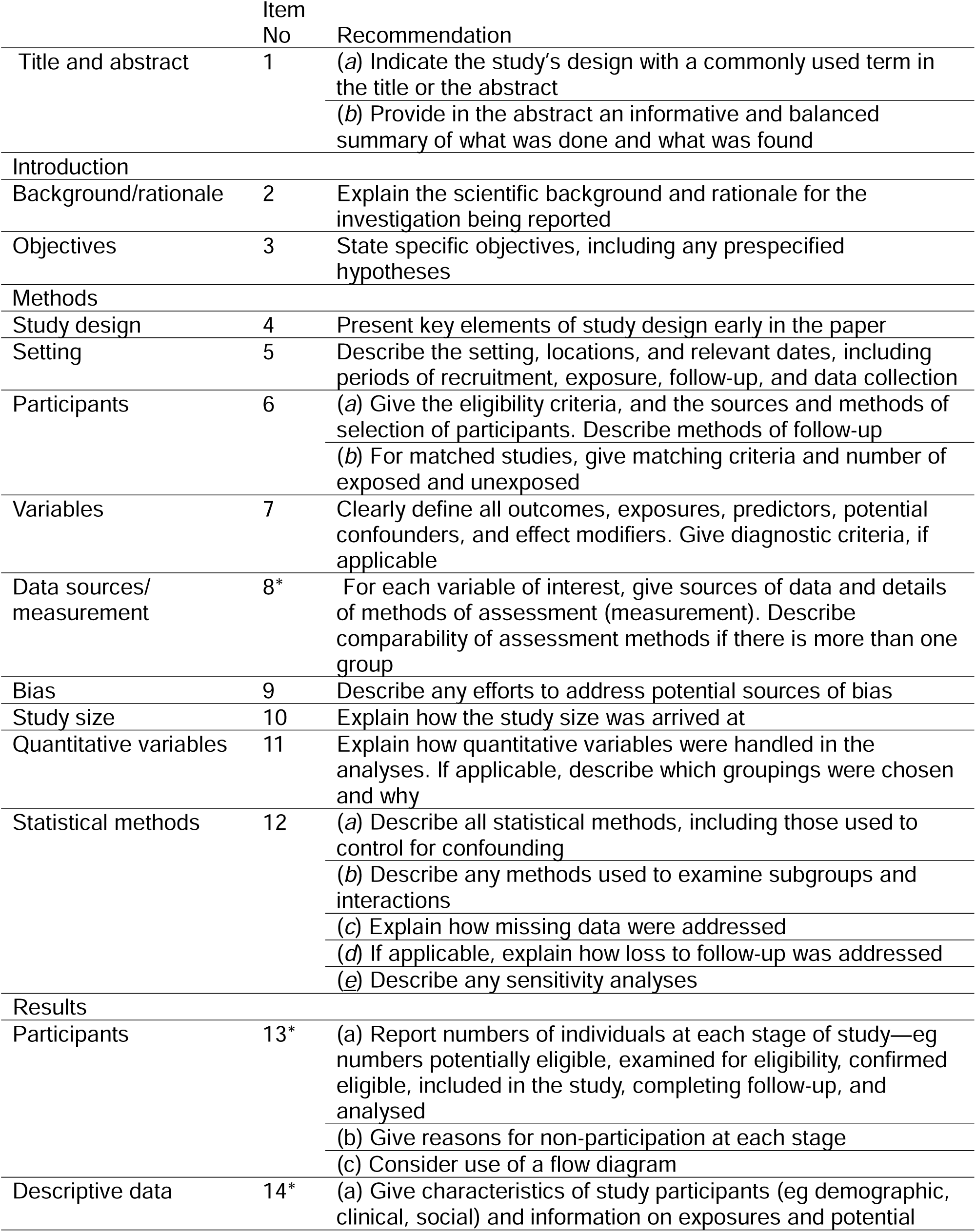
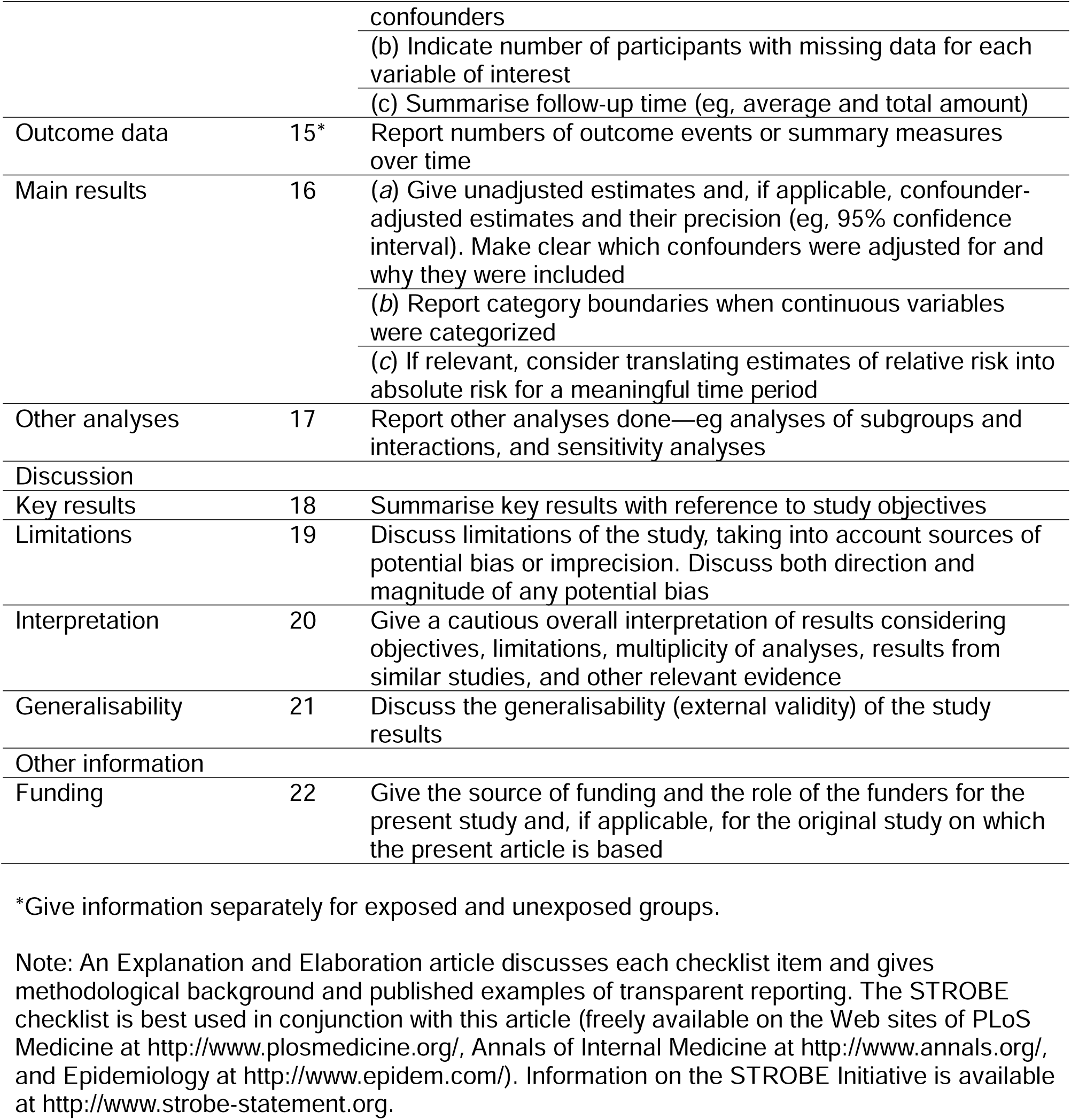

## Instrument to assess the credibility of effect modification analyses (ICEMAN) in a randomized controlled trial

*Version 1.1*

Consider the following important instructions informed by common misapplications of ICEMAN in studies using the instrument

- Complete a separate credibility assessment per each effect modifier (e.g., age, comorbidity, drug dose, etc.), outcome (e.g., mortality, stroke, duration of hospital stay), time-point (e.g., 3 months, 6 months), and effect measure (e.g. relative risk, risk difference).
- Do not apply ICEMAN if the interaction p-value is 0.1 or larger, i.e., provides very little statistical support for the existence of an effect modification (ICEMAN is designed to address the possible claim of an effect modification rather than the claim of no effect modification).
- Response options on the left indicate definitely or probably reduced credibility, response options on the right probably or definitely increased credibility
- Completely unclear should be interpreted as probably reduced credibility.
- To ensure transparency, provide a supporting comment under each question that provides a rationale for the rating.
- To ensure transparency, provide a copy of the completed ICEMAN instrument in the supplement of your article.

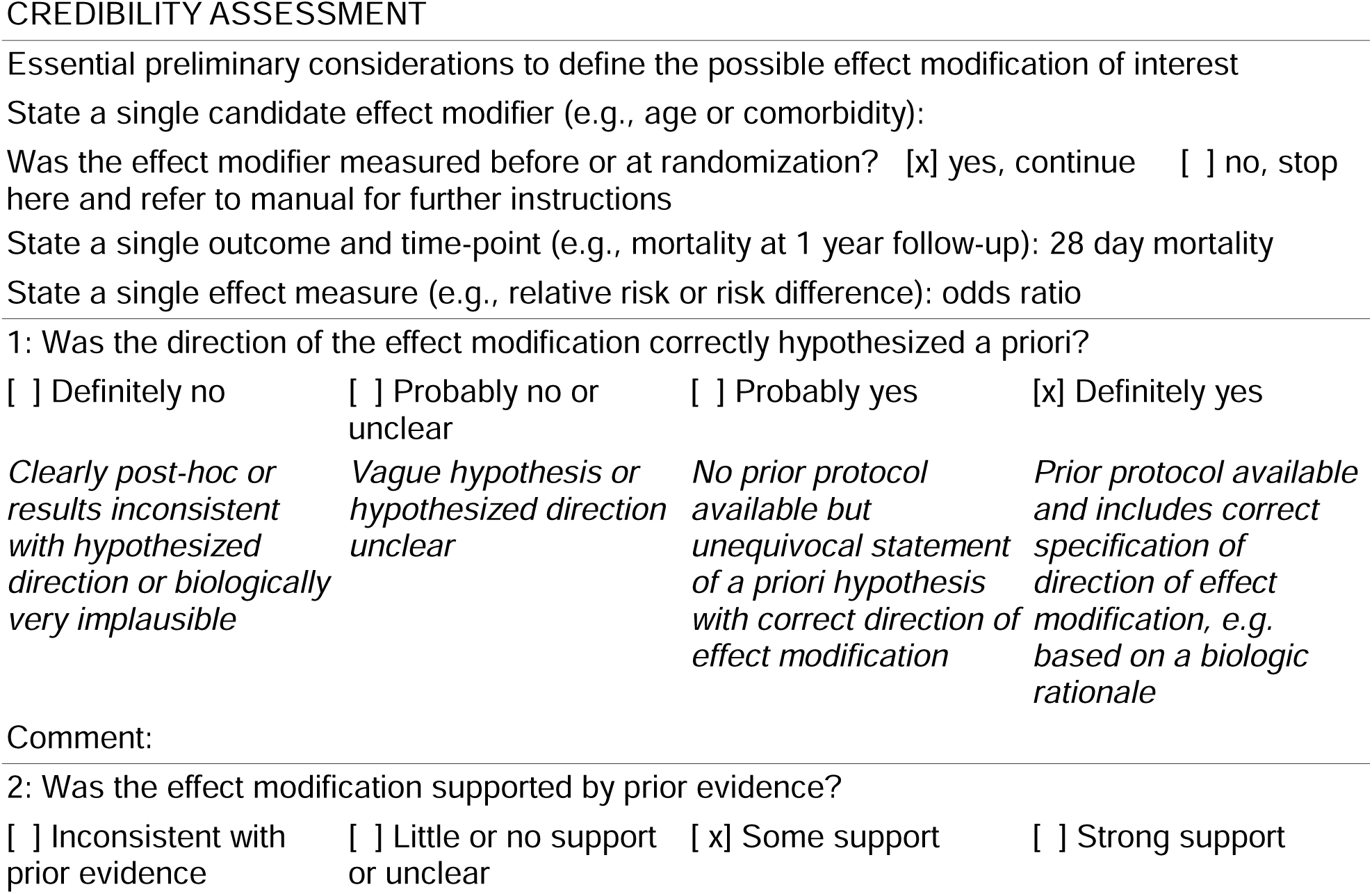
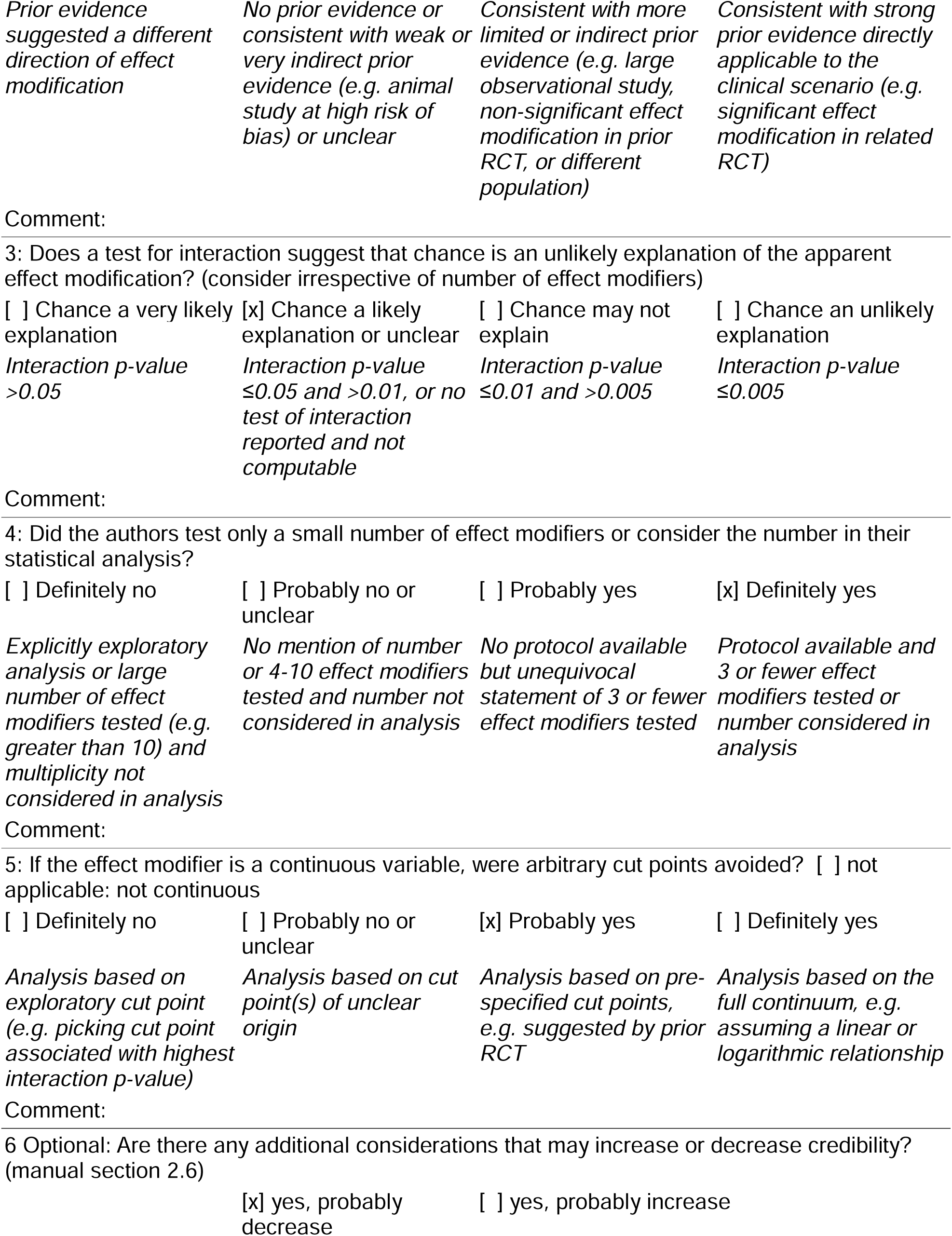
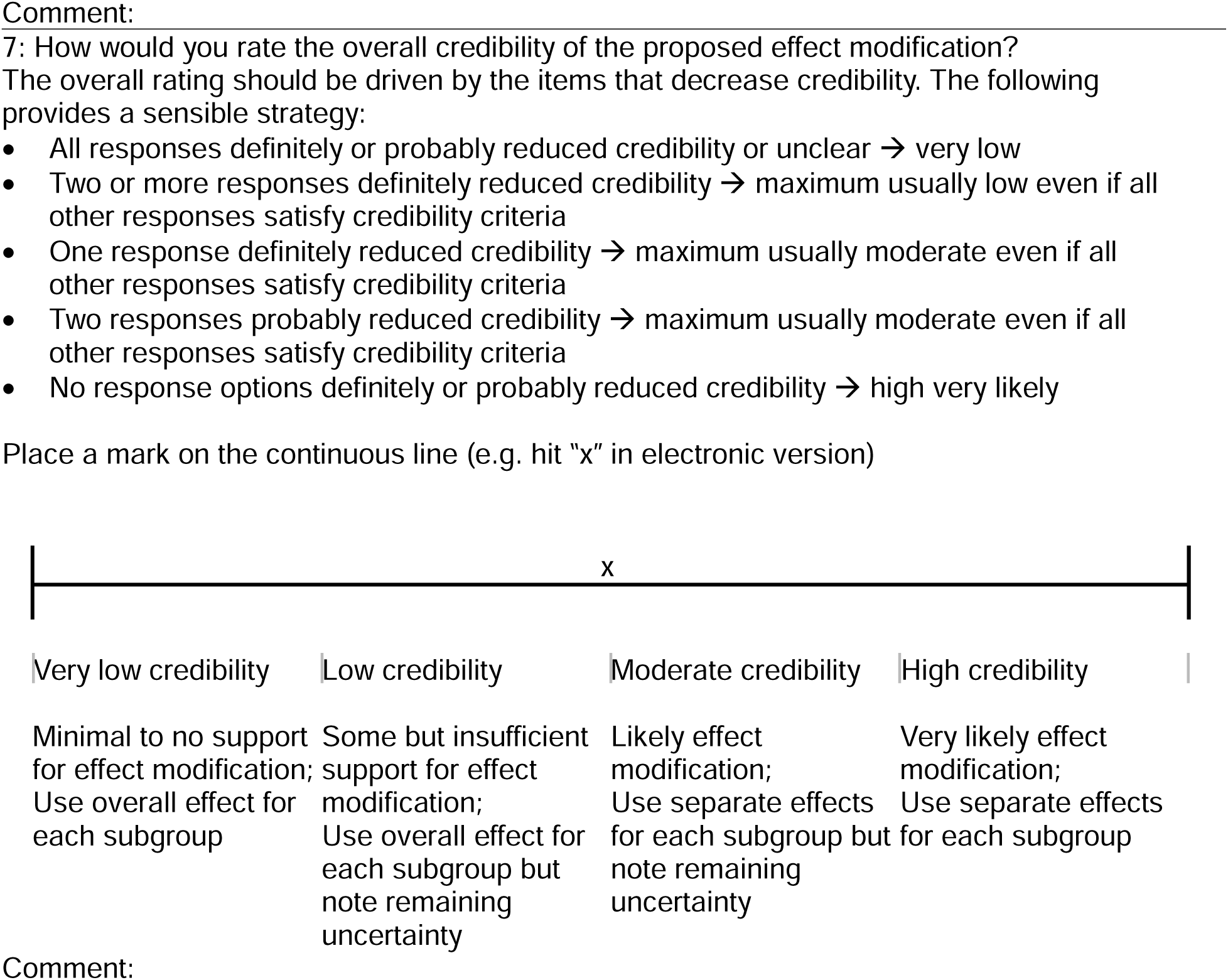

---

**Supplementary Table 1:** Emulated Target Trial.

|  | **Target Trial (RECOVERY)**<br>**\*Pragmatic version** | **Emulated Trial** |
| --- | --- | --- |
| Eligibility Criteria | Hospitalized with laboratory proven or suspect SARS-CoV-2 | Hospitalized with *acute hypoxic respiratory failure* and laboratory proven SARS-CoV-2 |
| *Cannot include restrictions based on post-baseline events* | Clinician discretion | Bone marrow transplant or solid organ transplant recipient<br>Acute myelogenous leukemia not in remission<br>Active treatment for malignancy<br>CAR-T therapy<br>Primary immunodeficiency<br>Advanced HIV or AIDS<br>Chronic corticosteroid use (defined as 20 mg or greater of prednisone or equivalent daily for greater than 14 days)<br>Receipt of other immune suppressing biologic agents<br>Other indication for corticosteroids (COPD exacerbation, adrenal insufficiency, severe shock) |
| Treatment Strategies | 6 mg of dexamethasone daily for 10 days or until hospital discharge | Glucocorticoid equivalent of 6 mg of dexamethasone daily for the shorter of 10 days or until hospital discharge |
| Assignment procedures | Randomized | Clinician discretion, stabilized inverse probability of treatment weighting |
| Follow-up period | 28 days | 28 days |
| Primary Outcome | Mortality | Mortality |
| Secondary Outcomes | Hospital Discharge<br>Composite of IMV/Death | Hospital Discharge<br>Composite of IMV/Death |
| Causal Contrast of Interest | Risk Ratio | Odds Ratio |
| *Per protocol effect* |  | Defined by med_admin exposure to corticosteroids |
| Analysis Plan |  | IPTW |

**Supplementary Table 2:**
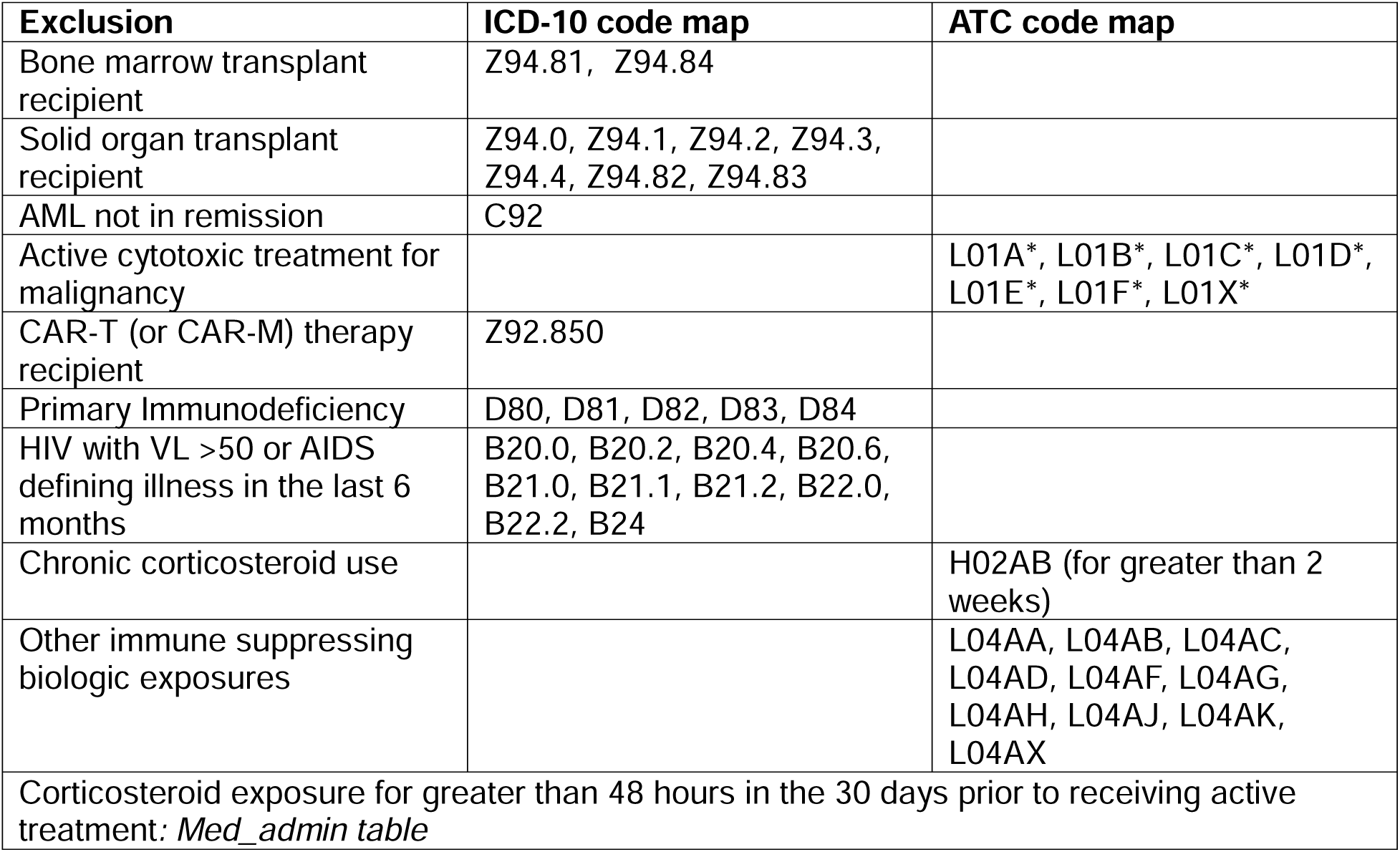
Exclusion Criteria.

**Supplementary Table 3:**
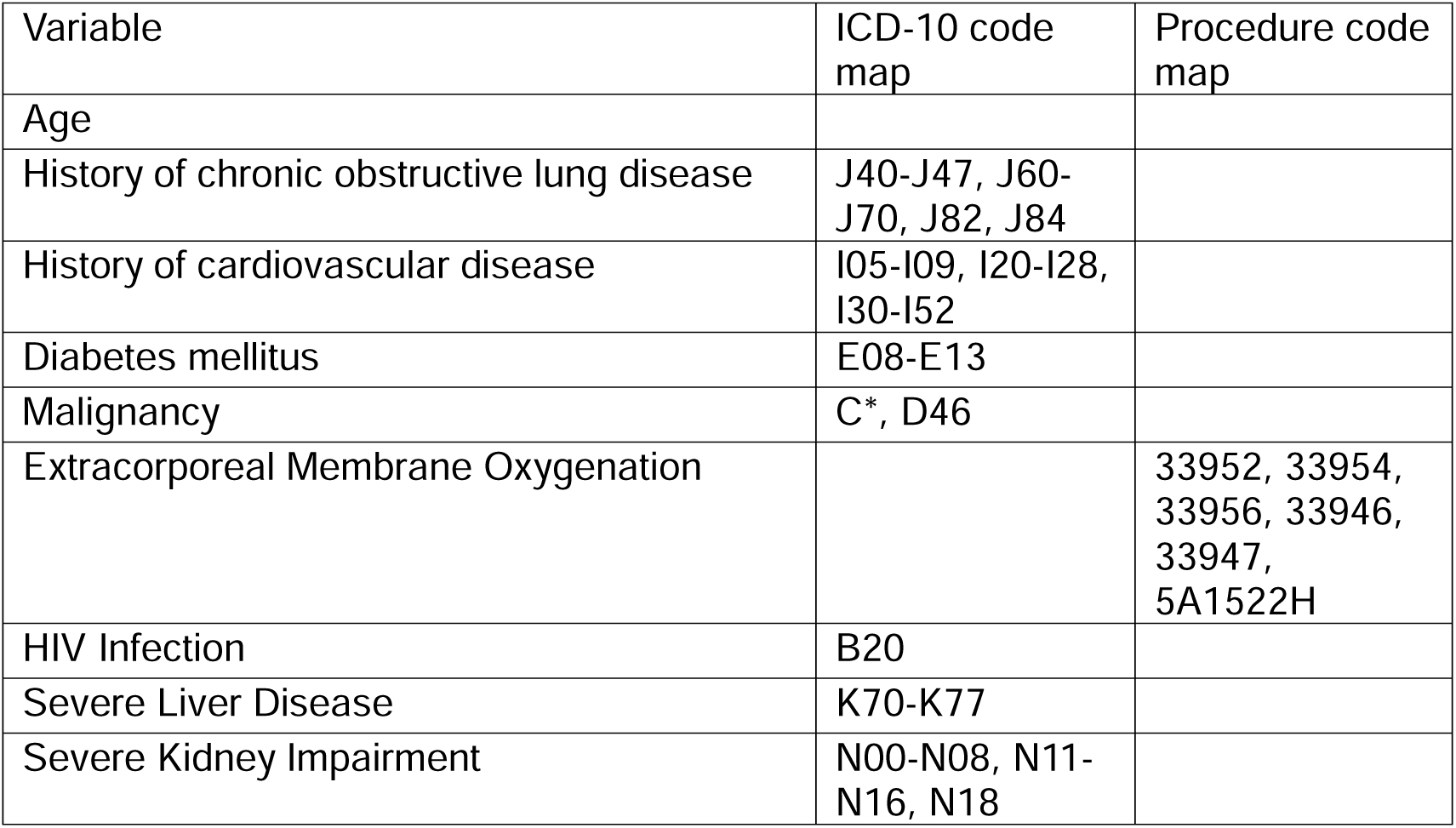
IPTW Specification.

| Variable | ICD-10 code map | Procedure code map |
| --- | --- | --- |
| Age |  |  |
| History of chronic obstructive lung disease | J40-J47, J60-J70, J82, J84 |  |
| History of cardiovascular disease | I05-I09, I20-I28, I30-I52 |  |
| Diabetes mellitus | E08-E13 |  |
| Malignancy | C*, D46 |  |
| Extracorporeal Membrane Oxygenation |  | 33952, 33954, 33956, 33946, 33947, 5A1522H |
| HIV Infection | B20 |  |
| Severe Liver Disease | K70-K77 |  |
| Severe Kidney Impairment | N00-N08, N11-N16, N18 |  |

**Supplementary Figure 1:**
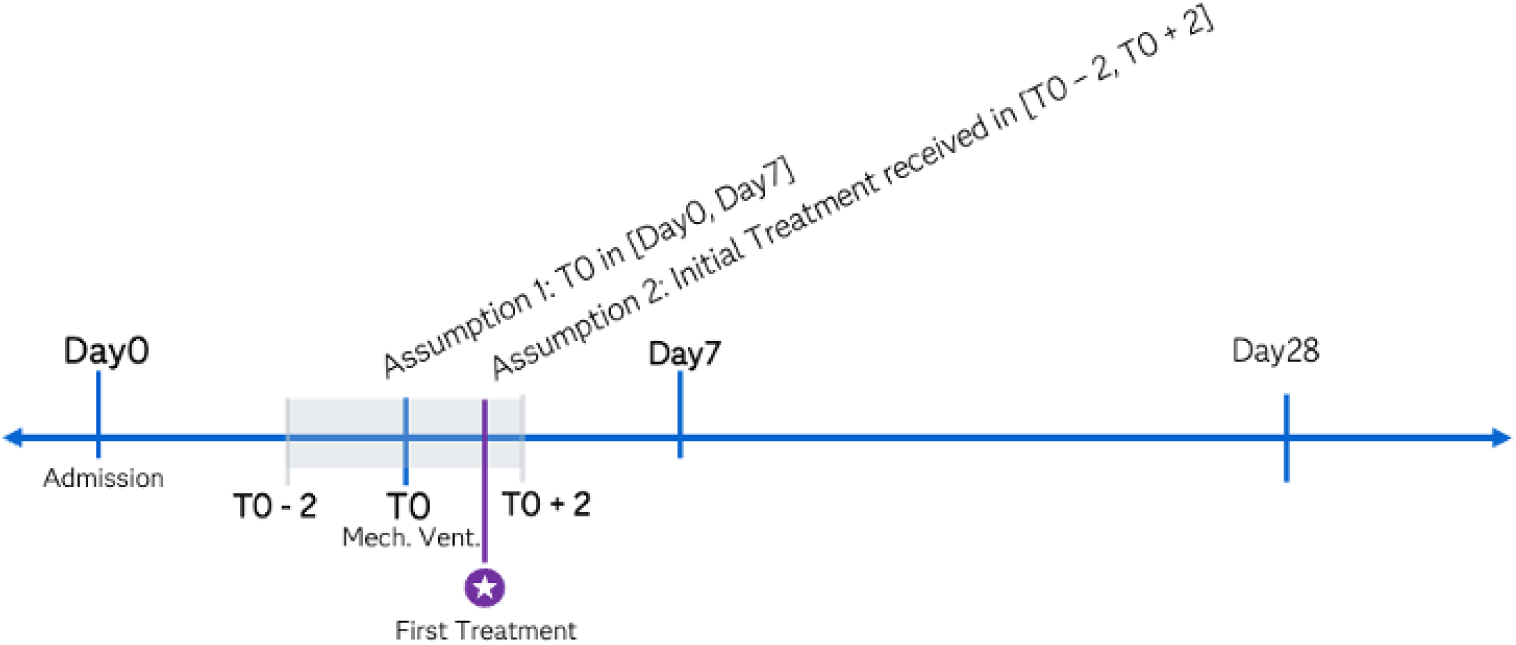
Sample timeline of patients assigned to the treatment arm.

**Supplementary Figure 2.**
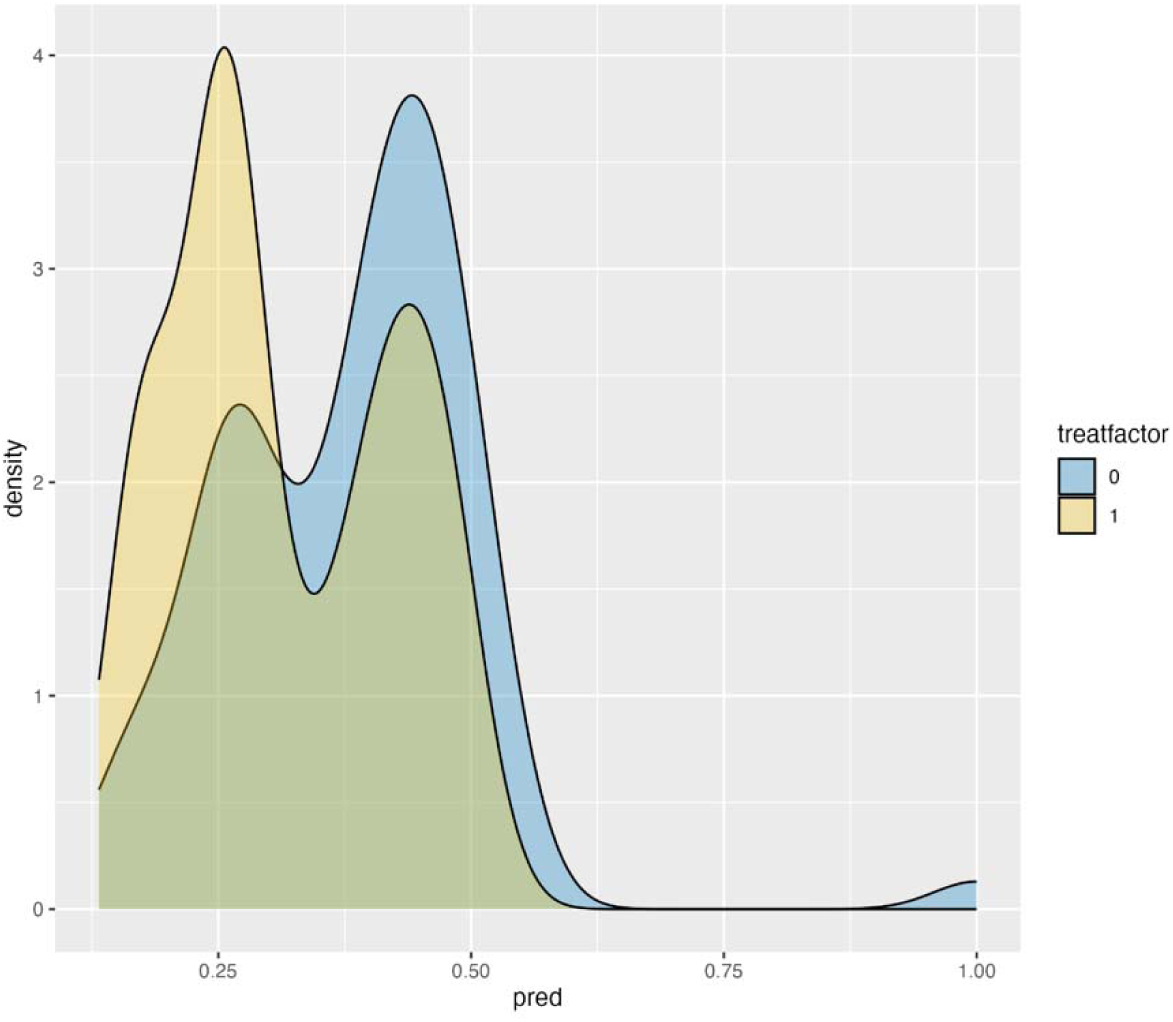
Distribution curves of IPTW scores between exposed and unexposed groups to assess for region of overlap

**Supplementary Figure 3.**
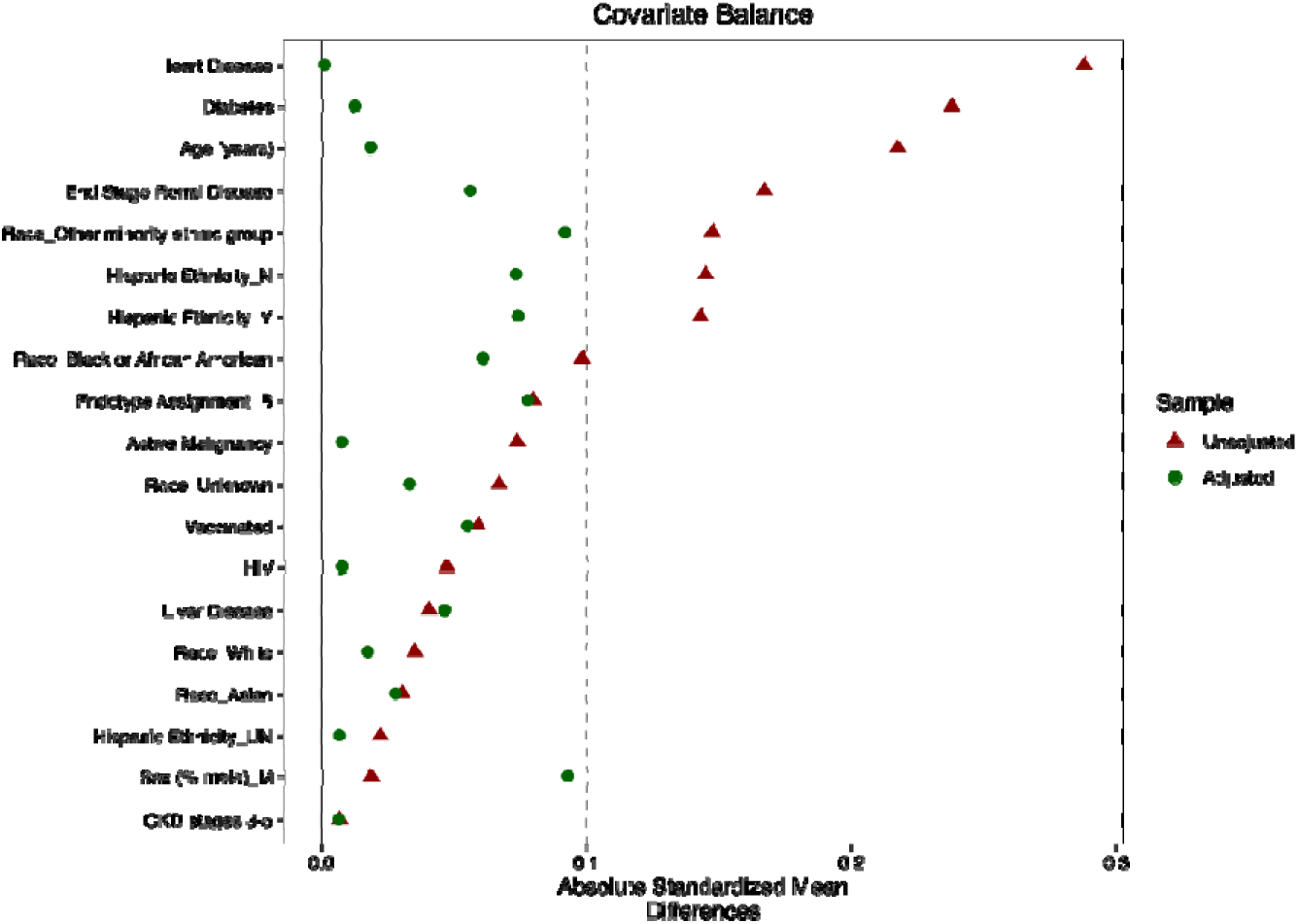
Love plot comparing balance of covariate in cohort population and weighted population

**Supplemental Figure 4:**
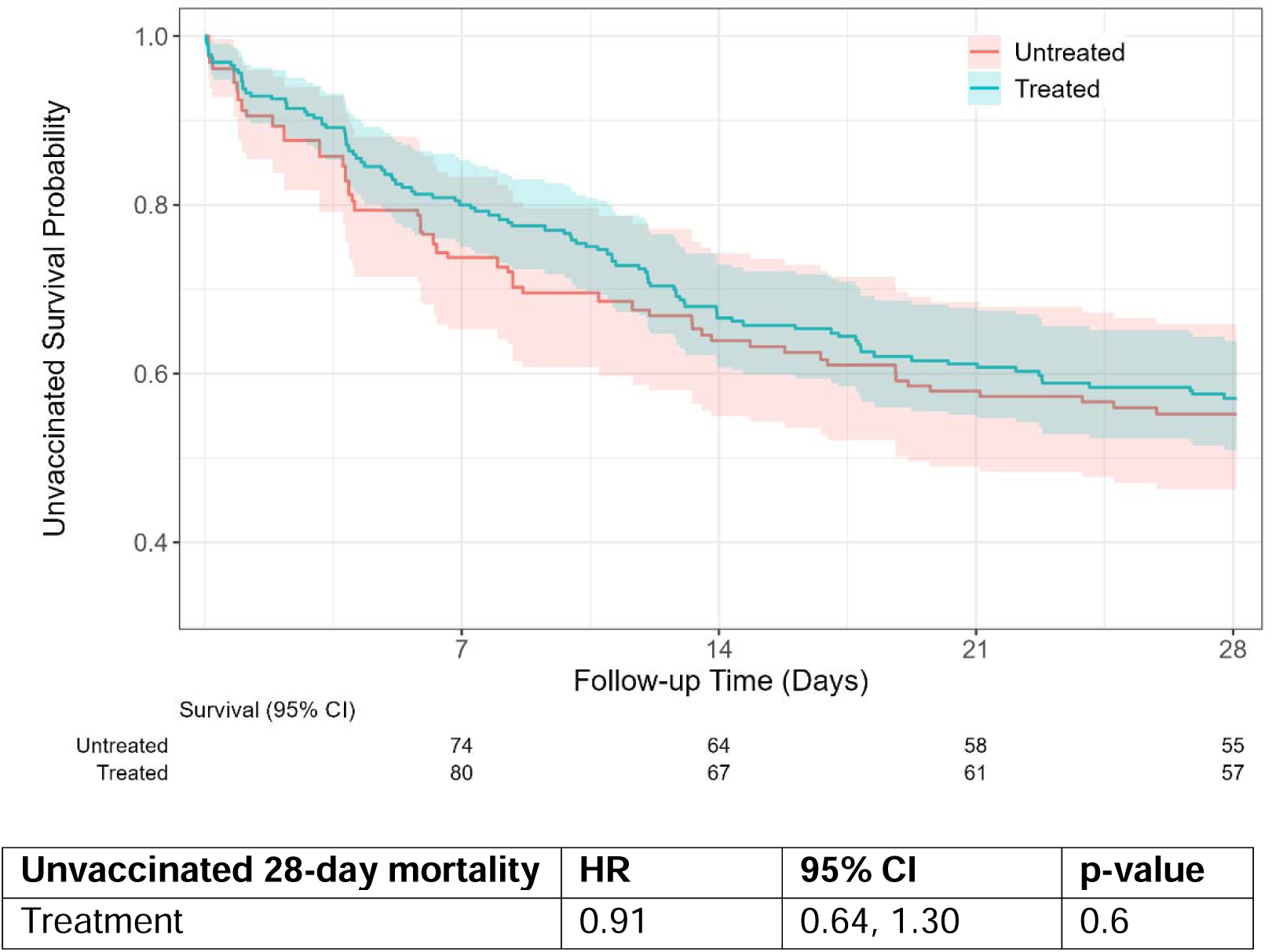
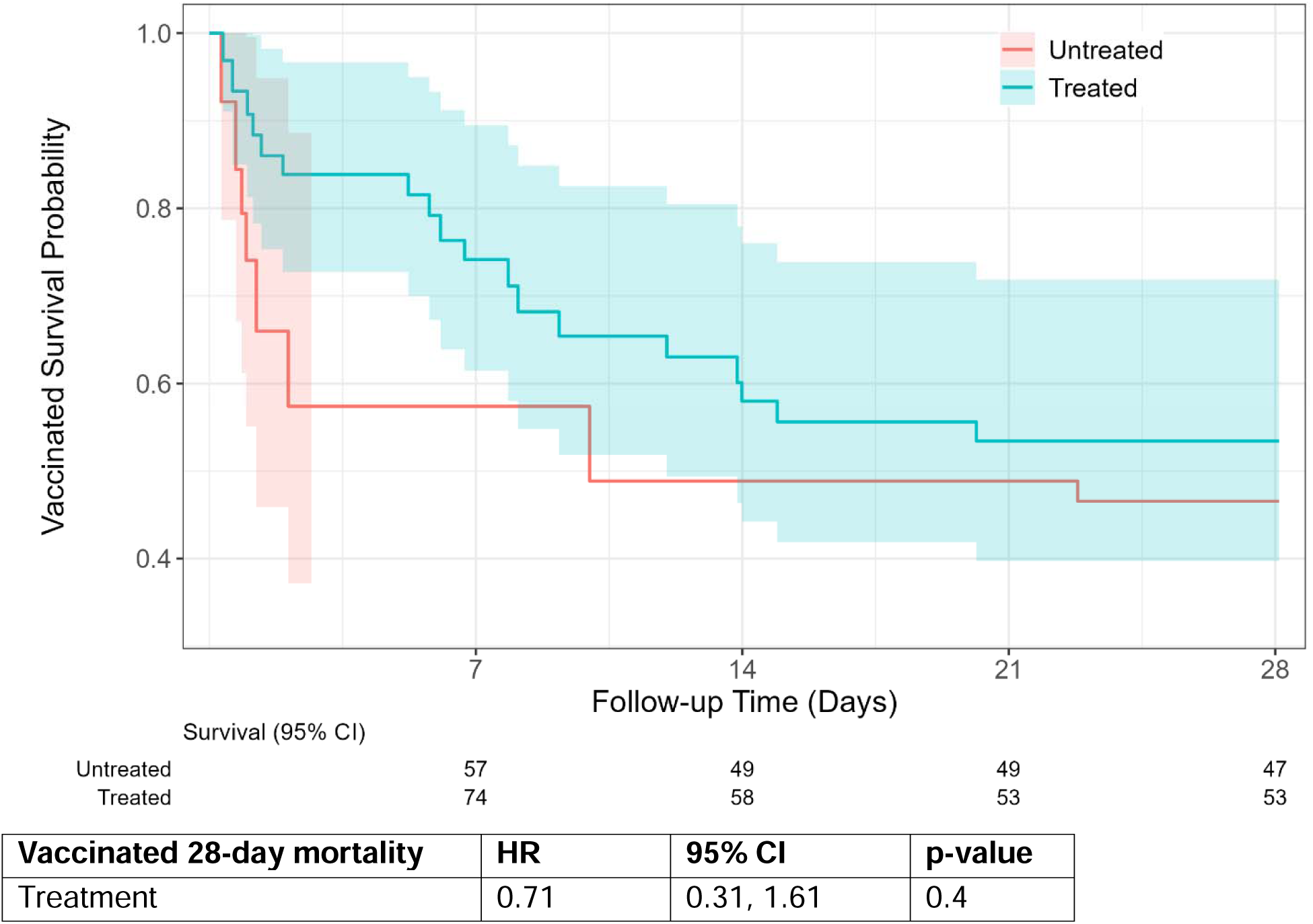
Corticosteroid effect by vaccine status on 28-day mortality

